# Persistent Inequities in Intravenous Thrombolysis for Acute Ischemic Stroke in the United States: Results from the Nationwide Inpatient Sample

**DOI:** 10.1101/2023.10.09.23296783

**Authors:** Philip Sun, Ling Zheng, Michelle Lin, Steven Cen, Gmerice Hammond, Karen E. Joynt Maddox, May Kim-Tenser, Nerses Sanossian, William Mack, Amytis Towfighi

**Author notes:** ADDRESS CORRESPONDENCE TO: Philip Young-woo Sun, MD Department of Neurology, David Geffen School of Medicine at University of California – Los Angeles 710 Westwood Plaza, Rm 1-240, Los Angeles, CA 90095.

## Abstract

**Background:** Despite its approval for use in acute ischemic stroke (AIS) >25 years ago, intravenous thrombolysis (IVT) remains underutilized, with inequities by age, sex, race/ethnicity, and geography. Little is known about IVT rates by insurance status. We aimed to assess temporal trends in the inequities in IVT use.

**Methods:** We assessed trends from 2002 to 2015 in IVT for AIS in the Nationwide Inpatient Sample by sex, age, race/ethnicity, hospital location/teaching status, and insurance, using survey*-* weighted logistic regression, adjusting for sociodemographics, comorbidities, and hospital characteristics. We calculated odds ratios for IVT for each category in 2002-2008 (*Period 1*) and 2009-2015 (*Period 2*).

**Results:** Among AIS patients (weighted N=6,694,081), IVT increased from 1.0% in 2002 to 6.8% in 2015 (adjusted annual relative ratio (AARR) 1.15, 95% CI 1.14-1.16). Individuals ≥85 years had the most pronounced increase from 2002 to 2015 (AARR 1.18, 1.17-1.19), but were less likely to receive IVT compared to those aged 18-44 years in both Period 1 (adjusted odds ratio (aOR) 0.23, 0.21-0.26) and Period 2 (aOR 0.36, 0.34-0.38). Women were less likely than men to receive IVT, but the disparity narrowed over time (Period 1 aOR 0.81, 0.78-0.84, Period 2 aOR 0.94, 0.92-0.97). Inequities in IVT by race/ethnicity resolved for Hispanic individuals in Period 2 but not for Black individuals (Period 2 aOR 0.81, 0.78-0.85). The disparity in IVT for Medicare patients, compared to privately insured patients, lessened over time (Period 1 aOR 0.59, 0.56-0.52, Period 2 aOR 0.75, 0.72-0.77). Patients treated in rural hospitals were less likely to receive IVT than those treated in urban hospitals; a more dramatic increase in urban areas widened the inequity (Period 2 urban non-teaching vs. rural aOR 2.58, 2.33-2.85, urban teaching vs. rural aOR 3.90, 3.55-4.28).

**Conclusion:** From 2002 through 2015, IVT for AIS increased among adults. Despite encouraging trends, only 1 in 15 AIS patients received IVT and persistent inequities remained for Black individuals, women, government-insured, and those treated in rural areas, highlighting the need for intensified efforts at addressing inequities.

## Introduction

Administration of intravenous thrombolysis (IVT) with recombinant tissue plasminogen activator in appropriately selected patients with acute ischemic stroke (AIS) is associated with improved mortality and functional outcomes^1,2^. With expansion of evidence-based systems of care, such as primary and comprehensive stroke center designation, IVT use has become more widespread over time in the United States,^3–6^ though it remains significantly underused in eligible populations.

Additionally, marked inequities remain by sex^7^, age^3^, race/ethnicity^7,8^, and geographic location^4,6,9^. For example, women are less likely to receive IVT than men, and individuals admitted to rural hospitals are less likely to receive IVT than those admitted to urban hospitals^3,7^. While some sociodemographic inequities in IVT use have improved over the past few decades (e.g. use of IVT in individuals ≥85 years^3^ from 2005 to 2010), others remain unresolved or have worsened (e.g. for women from 2007 to 2011^7^, for Black and Hispanic individuals from 2004 to 2010^8^, for all non-White individuals from 2007 to 2011^7^ and for people living in rural areas from 2000 to 2010^4^ and 2012 to 2017^9^).

Furthermore, although one could postulate that use of evidence-based protocols would reduce disparities, a recent study revealed that presentation to a primary stroke center enhanced rate of IVT use overall but did not alleviate racial disparities^8^.

Little is known about more recent temporal trends in IVT use by sex, race/ethnicity, age, and hospital location/teaching status. Additionally, to our knowledge, differences in IVT by insurance type have not been studied in the United States. Therefore, the aim of the study was to fill these gaps by evaluating recent temporal trends in IVT among individuals with AIS, stratified by age, race/ethnicity, sex, primary insurance, hospital teaching status, and urban/rural location using data from the National Inpatient Sample (NIS) from 2002 to 2015. We hypothesized an overall increase in IVT and reduction in inequities over the study period.

## Methods

### Population for Study

Data were obtained from the National Inpatient Sample (NIS), which was developed as part of the Healthcare Cost and Utilization Project (HCUP). Prior to 2012, the survey was designed to approximate a stratified 20% sample of all United States community hospitals (non-federal, short-term, general, and specialty hospitals) serving adults in the United States. From 2012, the sampling strategy transitioned to 20% of patient discharges from all United States community hospitals excluding rehabilitation and long-term acute care hospitals. The sampling strategy selected hospitals within states that have state inpatient databases according to defined strata based on ownership, bed size, teaching status, urban/rural location, and region^10^. All discharges from sampled hospitals for the calendar year were then selected for inclusion into NIS. To allow extrapolation for national estimates, both hospital and discharge weights are provided. Detailed information on the design of the NIS is available at http://www.hcup-us.ahrq.gov.

NIS captures discharge-level information on primary and secondary diagnoses and procedures, discharge vital status, and demographics on several million discharges per year. Data elements that could directly or indirectly identify individuals are excluded. The unit of analysis is the discharge rather than the individual; discharges are therefore all considered independent. A unique anonymous hospital identifier allows for linkage of discharge data to an NIS data set with hospital characteristics. To protect subject confidentiality, NIS data only provides hospital-specific identifiable information (e.g., hospital rurality, but not the rurality of patient residence).

We included all patients with a primary or secondary discharge diagnosis of stroke (International Classification of Diseases, Ninth Revision diagnosis codes [ICD-9-CM] 433.01, 433.11, 433.21, 433.31, 433.81, 433.91, 434.01, 434.91, 436) at the time of hospital admission, from January 2002 through September 2015. IVT administration was determined using ICD-9 procedure code 99.10.

We excluded patients with a diagnosis of acute myocardial infarction, pulmonary embolism, malignancy (solid tumor without metastasis, lymphoma, metastatic cancer), transferred to index hospital from another hospital, elective admissions, cases with missing race/ethnicity and/or sex, and enrollment in a clinical trial (ICD-9-CM code V70.7). Please refer to Supplemental Tables 1-3 for the full list of ICD-9 codes.

### Sociodemographic, Clinical, and Hospital Factors

Individuals were categorized into the following age groups: <18 years, 18-44 years, 45-64 years, 65-84 years, and ≥85 years. They were also categorized by sex (women/men), race/ethnicity (White, Black, Hispanic, Asian/Pacific Islander (API), Native American, other, missing), primary payer (Medicare, Medicaid, private, self-pay, no charge, other pay, missing), and hospital location/teaching status (urban teaching, urban non-teaching, rural). Race/ethnicity was determined from two HCUP administrative data elements of race and ethnicity. If the source supplied race and ethnicity in separate data elements, then ethnicity took precedence over race.

Records with missing race/ethnicity were placed into an independent category of “Missing Race” and were included in the analysis. Risk of mortality was determined by the Risk of Mortality Subclass Category^11^. Presence of the following comorbid conditions were assessed: hypertension, dyslipidemia, alcohol abuse, obesity, smoking history, coronary artery disease, atrial fibrillation, and Charlson Comorbidity Index^12^ (which consists of 17 comorbidities; Supplemental Table 3).

### Statistical Analyses

National trends were estimated following HCUP methodological standards (which adopted a design change in 2012), with appropriate trend weights. The observed yearly national IVT utilization from 2002 to quarter 3 of 2015 was estimated using proc surveyfreq in SAS. For the following demographic, comorbidities, and hospital factors, we conducted national estimations by year, as well as by IVT utilization status: sex, age, race/ethnicity, national quartile of household income by zip code, third-party payer, hospital region/teaching status, coronary artery disease, atrial fibrillation or flutter, hypertension, dyslipidemia, obesity, smoking, alcohol use, Charlson Comorbidity Index, and Risk of Mortality Subclass. Any factors with an observed association with both year and with IVT utilization in univariate analysis or trend analysis meaningfully in either clinical or epidemiological ways were considered as confounders that could impact the temporal trend effects of IVT. To systematically compare temporal trends, we divided the study period 2002-2015 into two periods – 2002-2008 (*Period 1*) and 2009-2015 (*Period 2*).

The temporal trend effect in IVT was tested using survey-weighted logistic models, adjusting for sex, age, race/ethnicity, national quartile of household income by zip code, insurance, hospital region, hospital location/teaching status, hypertension, diabetes, dyslipidemia, obesity, coronary artery disease, peripheral artery disease, atrial fibrillation, congestive heart failure, Charlson Comorbidity Index, smoking history, and alcohol history. In addition to the linear temporal trend, we compared first and second seven-year periods (*Period 1* vs. *Period 2*) adjusting for the same covariates. For temporal trends in subgroups, we used the survey-logistic procedure with subgroup categories in the DOMAIN statement (https://www.hcupus.ahrq.gov/reports/methods/2003_02.jsp#sas) to request statistics for the subpopulation. All data analyses were conducted using SAS (version 9.4; SAS Institute Inc, Cary, NC).

The study was considered exempt from institutional review board given the use of deidentified information. We followed the Strengthening the Reporting of Observational Studies in Epidemiology (STROBE) reporting guideline^13^.

## Results

Among patients admitted with a diagnosis of AIS (weighted n=6,694,081), approximately half were 65-84 years (50.5%), just under a quarter were 45-64 years (24.0%), and 21.5% were ≥85 years of age **(Table 1**). More than half of the study population were women (53.9%). The race/ethnic distribution was as follows: White (58.9%), Black (14.5%), Hispanic (6.6%), API (2.2%), and other (2.1%). Nearly two thirds of patients had comorbid hypertension (61.4%) and approximately one in four had diabetes (26.6%), coronary artery disease (23.9%), or atrial fibrillation (22.4%). Medicare was the most common insurance type (70.3%), followed by private (16.8%) and Medicaid (6.7%). The distribution by geography was as follows: South (41.0%), Midwest (22.0%), Northeast (18.5%) and West (18.5%).

**Table 1.**
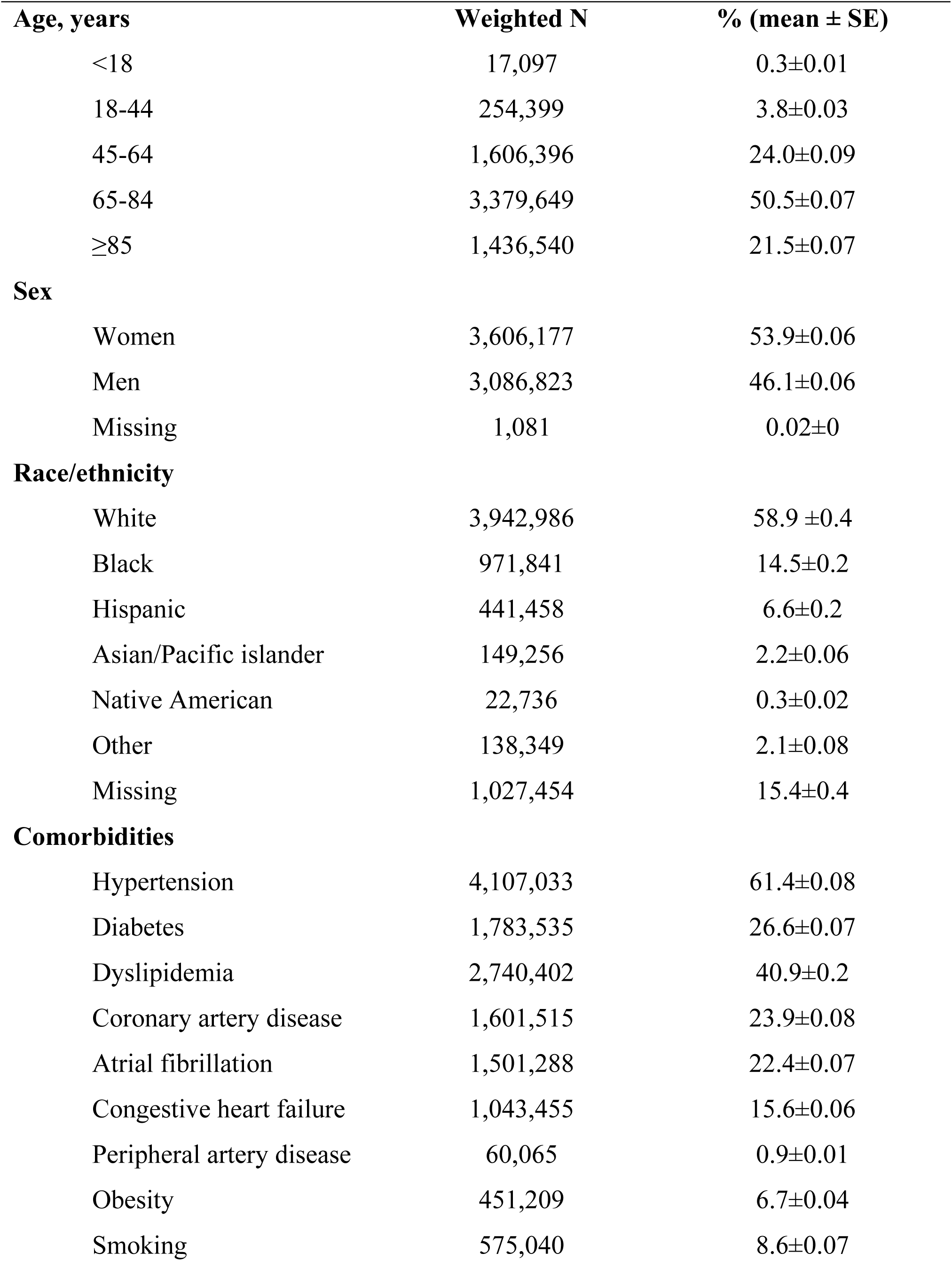
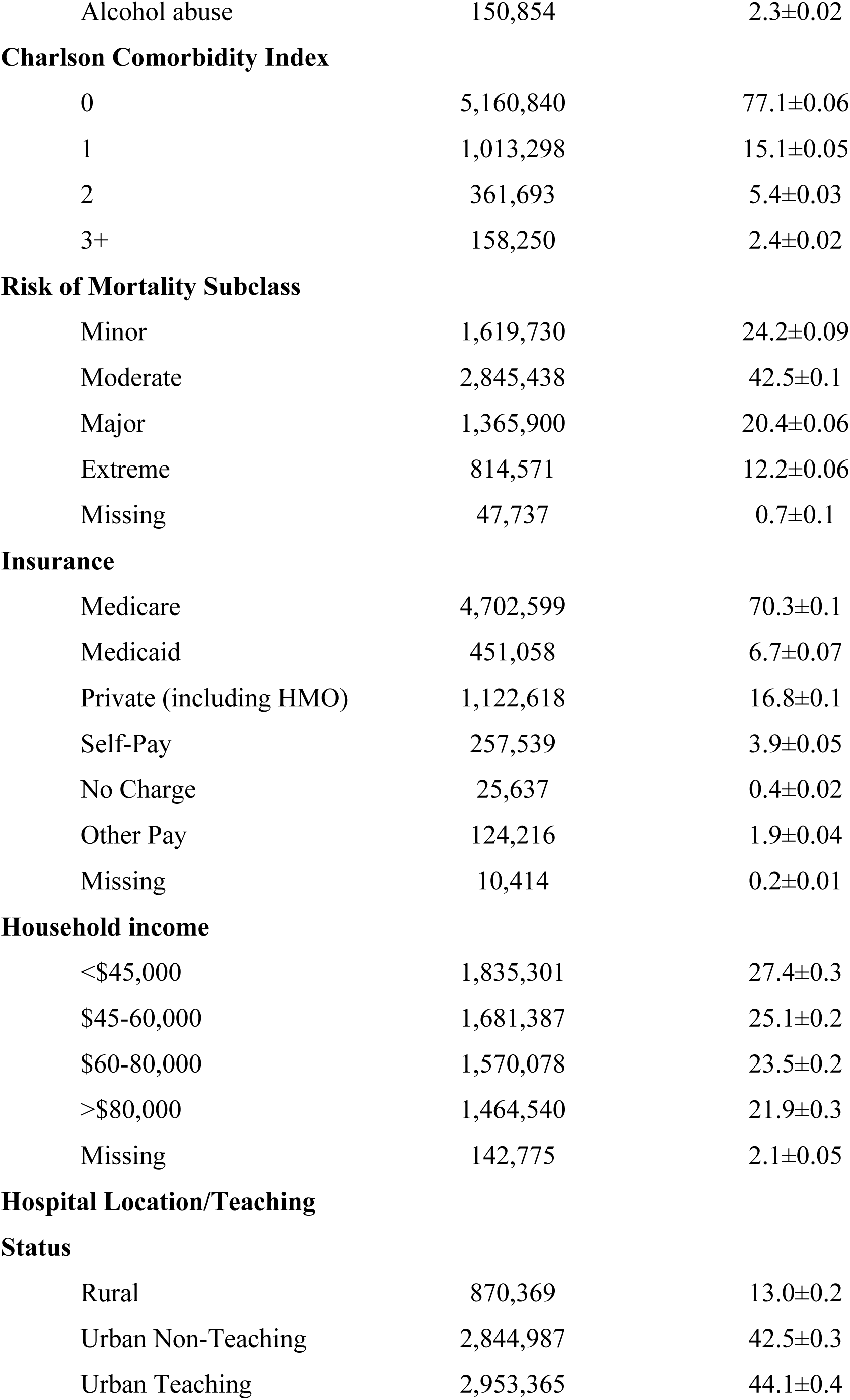
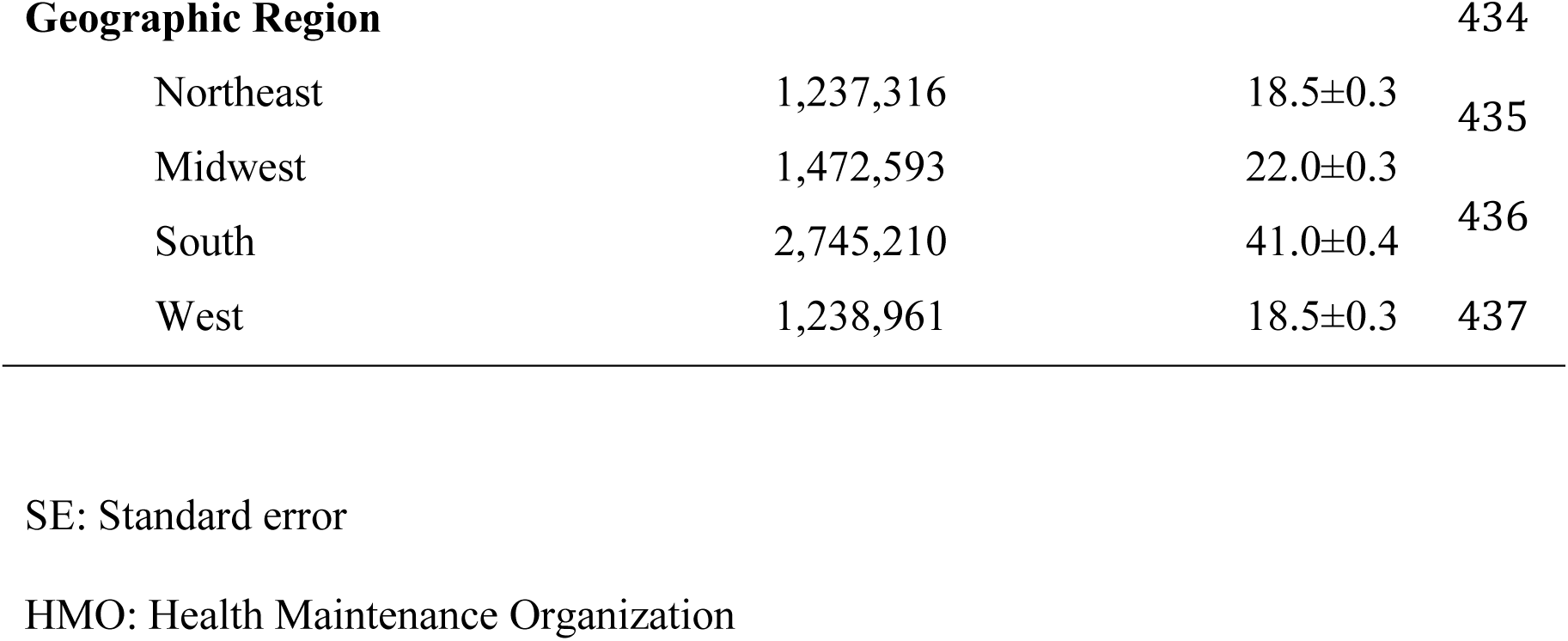
Patient and hospital characteristics of overall cohort (weighted N=6,694,081)

The overall use of IVT for AIS increased from 1.0% in 2002 to 6.8% in 2015, with an overall adjusted annual relative ratio (AARR) of 1.15 (95% CI 1.14-1.16). Individuals aged 18-44 years had the highest rate of IVT during the entire period starting in 2003 (**Figure 1**). Adults 18 years of age or greater had an increase in IVT over time, with those ≥85 years having the most pronounced increase (AARR of 1.18, CI 1.17-1.19; Period 2 vs. Period 1 adjusted odds ratio (aOR) of 3.66, CI 3.3-4.07) (**Table 2**). However, compared to those aged 18-44 years, those who were ≥85 years were still markedly less likely to receive IVT in both Period 1 (aOR 0.23, 95% CI 0.21-0.26) and Period 2 (aOR 0.36, 95% CI 0.34-0.38; **Figure 2A**). Minors (age <18) had the second lowest rate of IVT in Period 1 and the lowest in Period 2, and they were the only age group without a significant increase in IVT use over time (AARR 0.95, CI 0.88-1.03; Period 2 vs. Period 1 aOR of 0.94, CI 0.49, 1.77)(**Table 2**).

**Figure 1.**
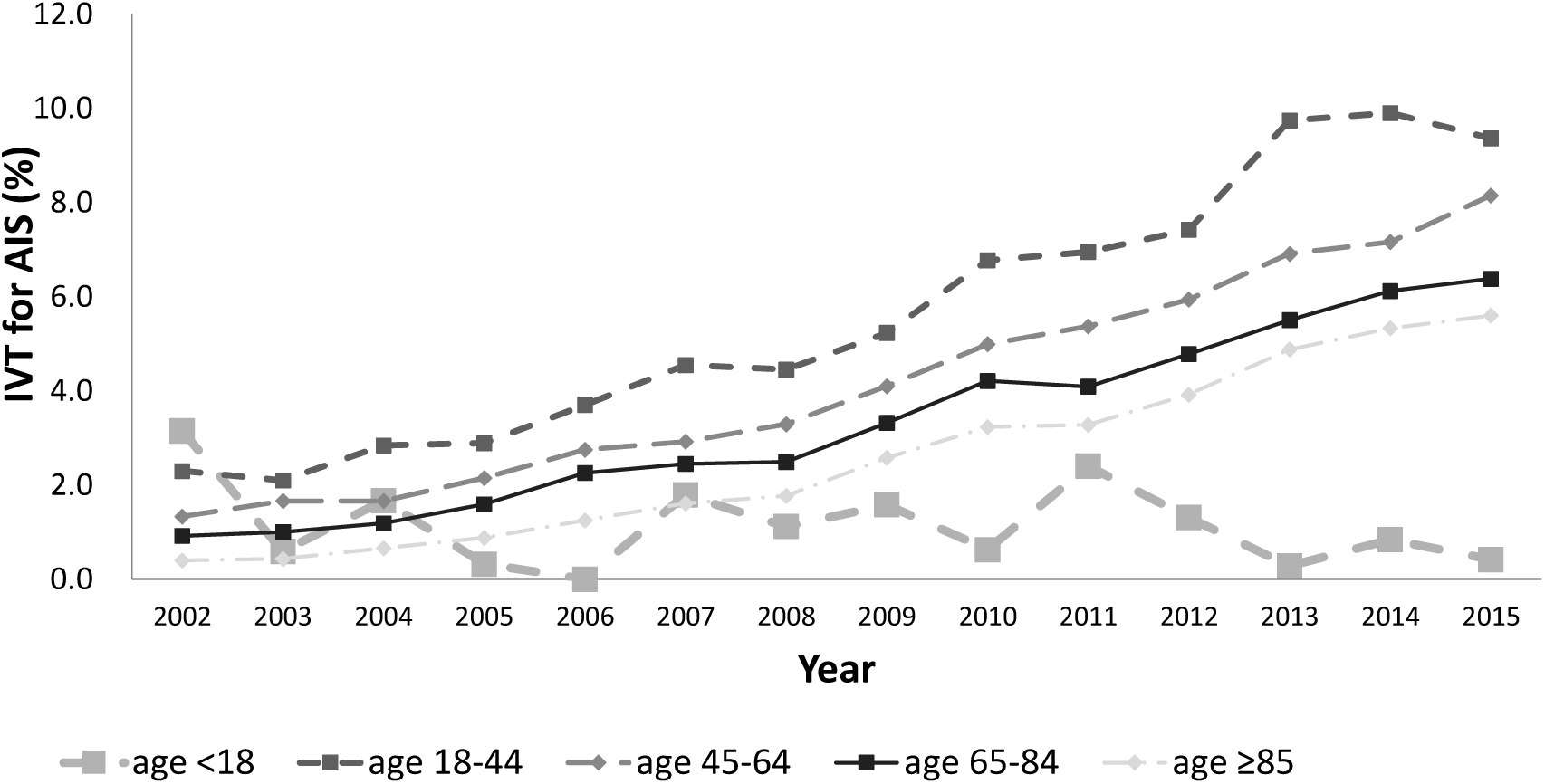
Proportion of acute ischemic stroke patients who received IVT by age from 2002 to 2015 (unadjusted)

**Figure 2A.**
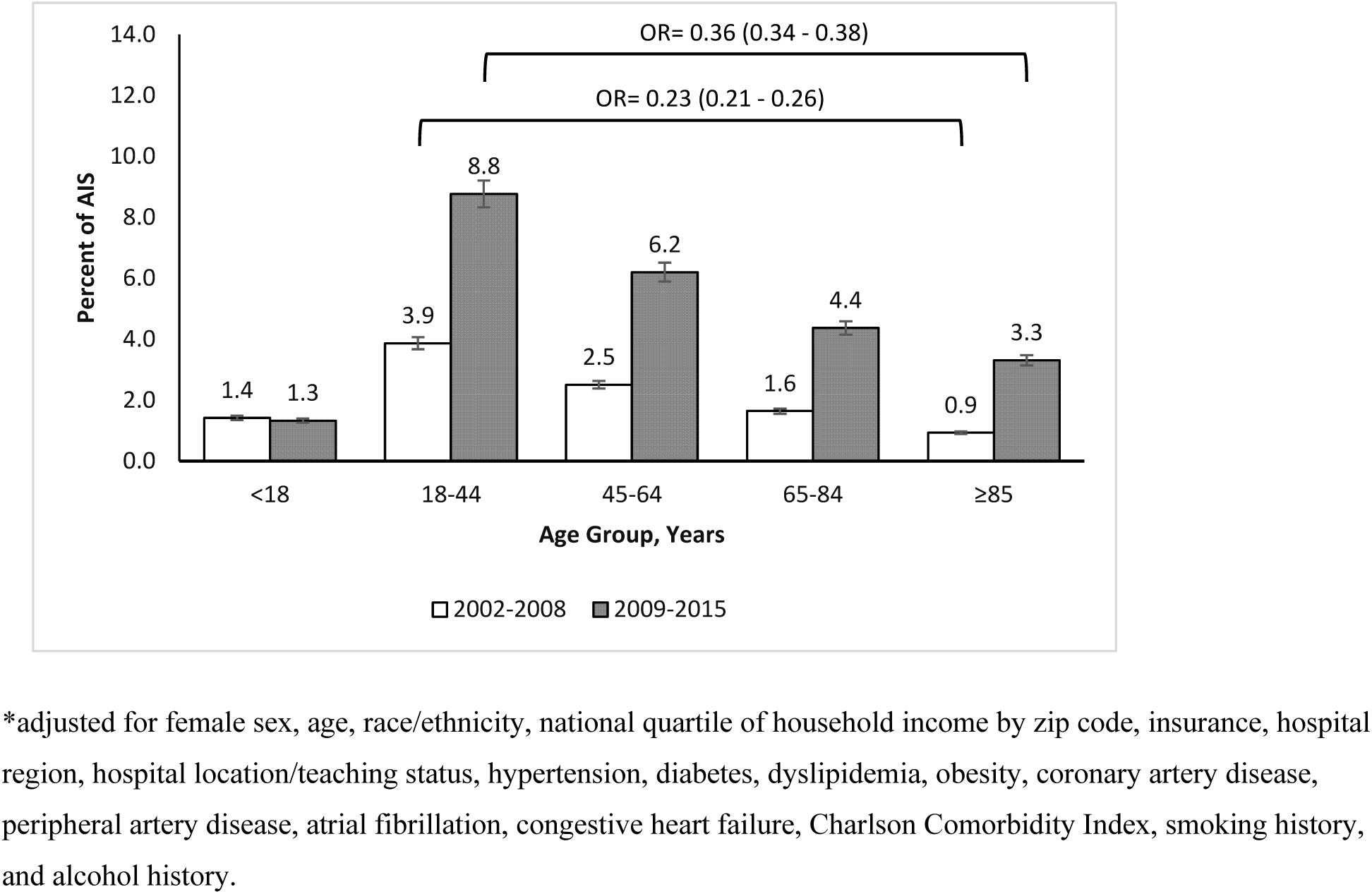
Dichotomized comparison of IVT use between *Period 1* vs. *Period 2* by age

**Table 2:**
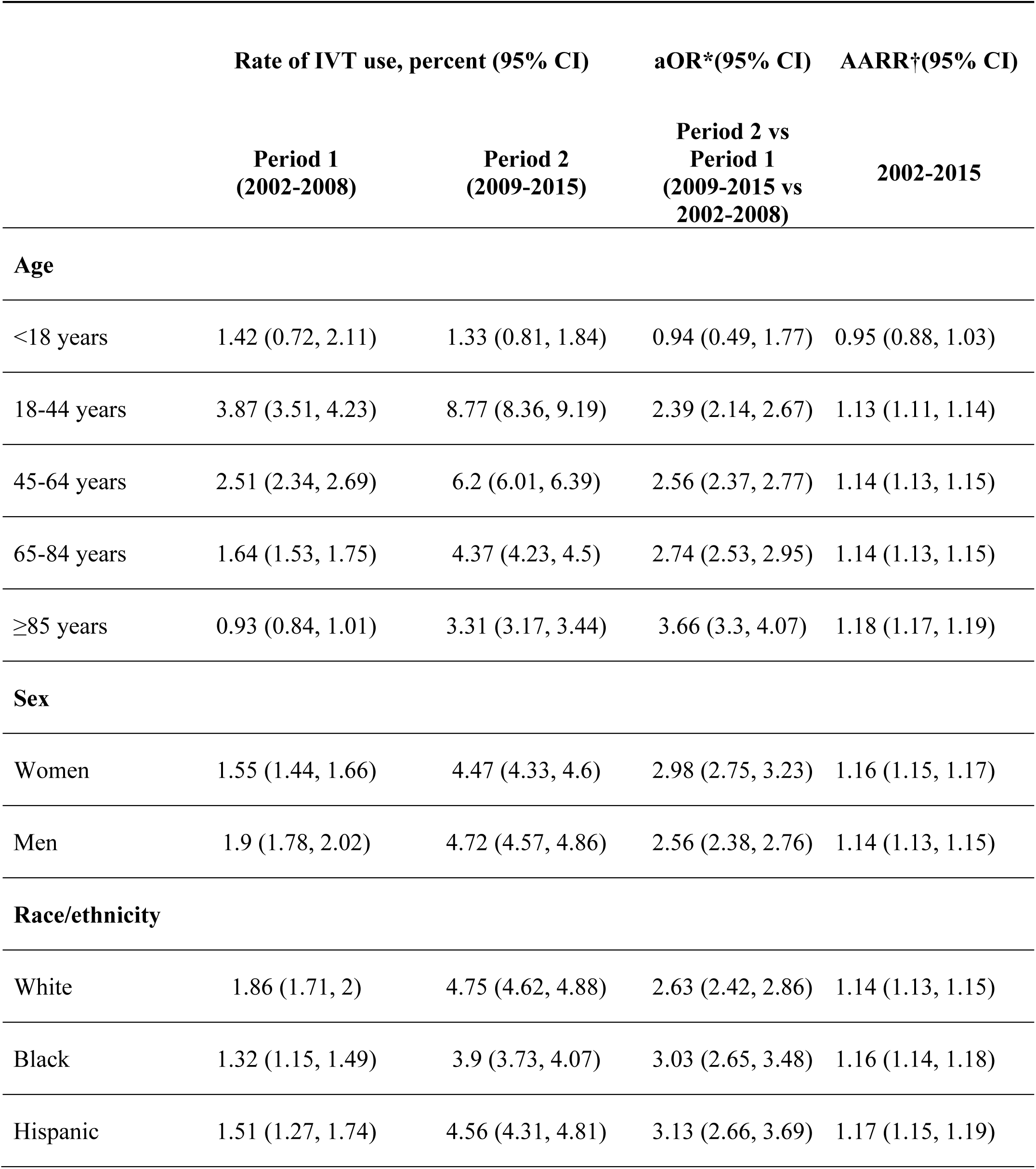
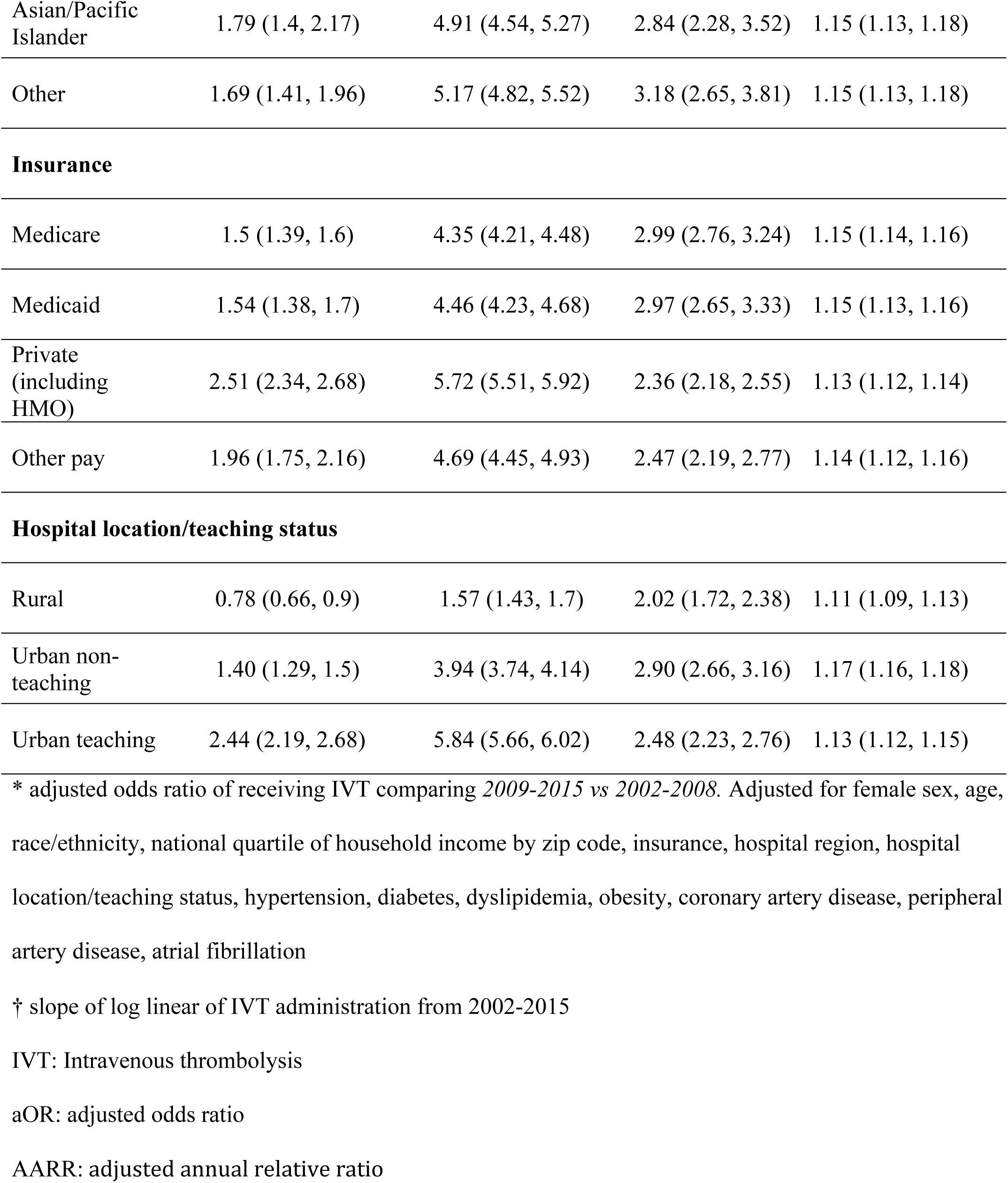
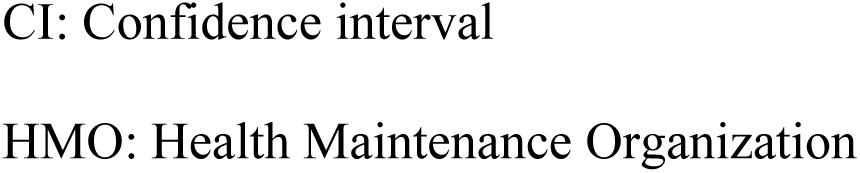
Rate of IVT from 2002 to 2015 by age groups, race/ethnicity, sex, insurance, and hospital location/teaching status.

Women were less likely than men to receive IVT in Period 1 (aOR 0.81, 95% CI 0.78-0.84; **Figure 2B**); this inequity narrowed such that women remained slightly less likely than men to receive IVT in Period 2 (aOR 0.94, 95% CI 0.92-0.97; **Figure 2B**). Compared to Period 1, women in Period 2 were about three times more likely than to receive IVT (aOR 2.98, 95% CI 2.75-3.23)(**Table 2**).

**Figure 2B.**
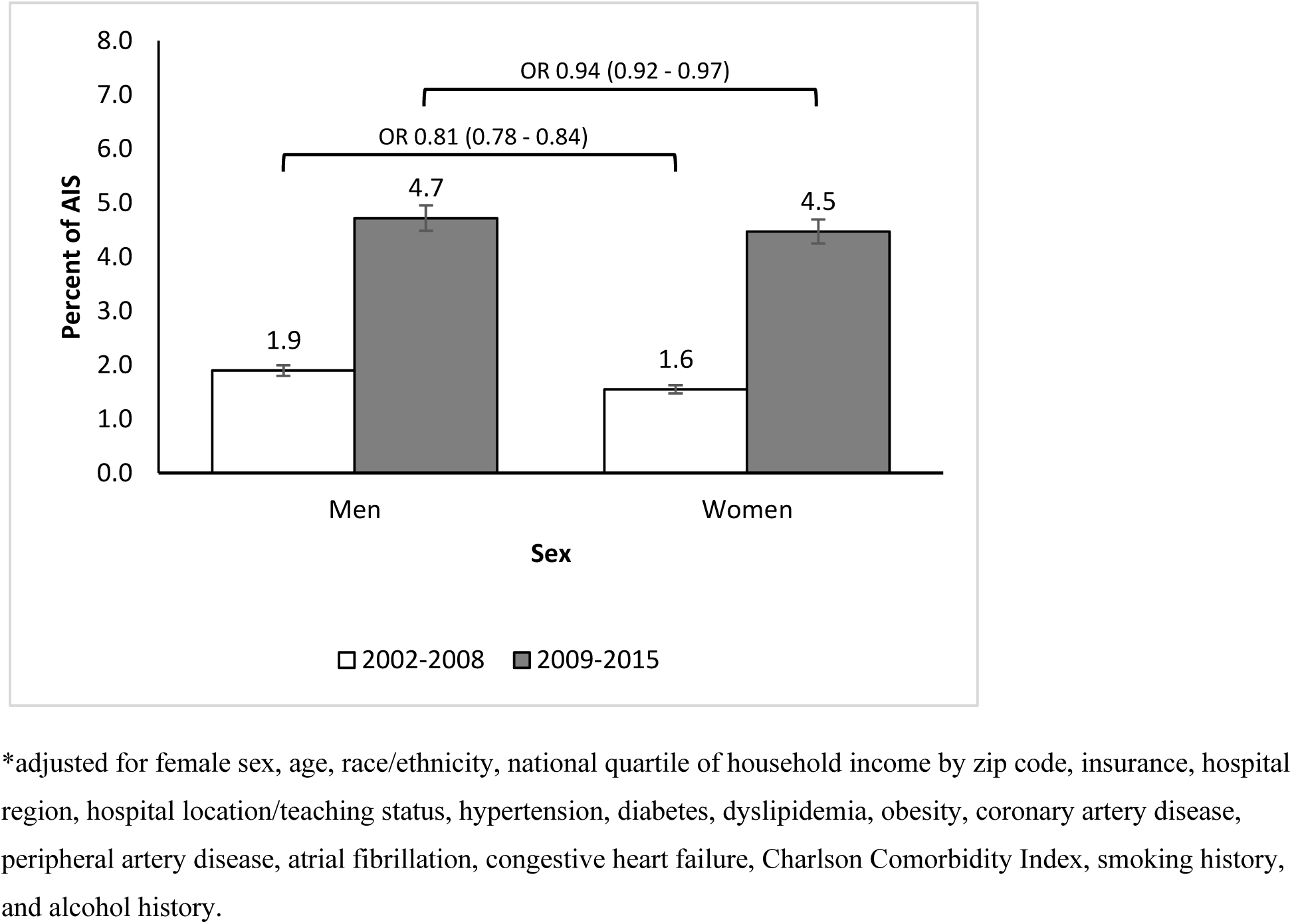
Dichotomized comparison of IVT use between *Period 1* vs. *Period 2* by sex

Across all races/ethnicities, IVT rates increase; therefore, individuals were more likely to receive IVT in Period 2 vs. Period 1 (White population OR 2.63, 95% CI 2.42-2.86; Black population aOR 3.03, 95% CI 2.65-3.48; Hispanic population aOR 3.13, 95% CI 2.66-3.69; Asian/Pacific Islander population aOR 2.84, 95% CI 2.28-3.52; other race/ethnicity aOR 3.18, 95% CI 2.65-3.81) (**Table 2**). In Period 1, Hispanic individuals were less likely than their non-Hispanic White counterparts to receive IVT (aOR 0.81, 95% CI 0.69-0.94; **Figure 2C**), but the inequity resolved in Period 2 (aOR 0.96, 95% CI 0.91-1.02; **Figure 2C**). Inequities in IVT for Black compared to White individuals with stroke improved without resolving. In Period 1, compared to White individuals, Black individuals were less likely to receive IVT (Period 1 aOR 0.71, 95% CI 0.63-0.79; Period 2 aOR 0.81, 95% CI 0.78-0.85; **Figure 2C**). Compared to Period 1, Black individuals in Period 2 were three times more likely to receive IVT (aOR 3.03; 95% CI 2.65-3.48; **Table 2**).

**Figure 2C.**
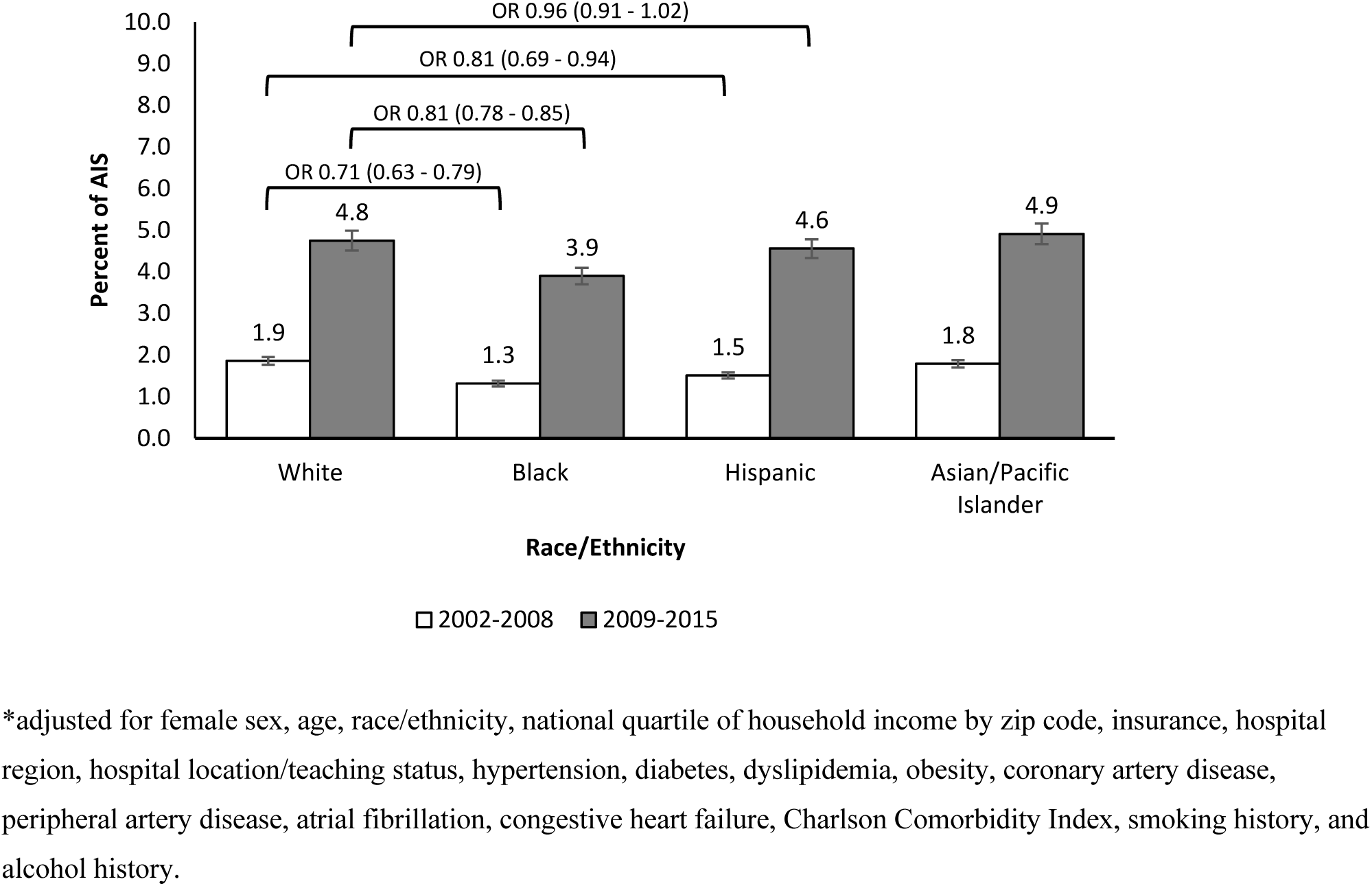
Dichotomized comparison of IVT use between *Period 1* vs. *Period 2* by race/ethnicity

In Period 1, compared to those who were privately insured, individuals with Medicare were less likely to receive IVT, even accounting for age (aOR 0.59, 95% CI 0.56-0.62; **Figure 2D**). This disparity improved but persisted in Period 2 (aOR 0.75, 95% CI 0.72-0.77; **Figure 2D**). Compared to Period 1, those with Medicare were three times more likely to receive IVT in Period 2 (aOR 2.99, 95% CI 2.76-3.24; **Table 2**). Those with Medicaid insurance had a similar trend of marginal improvement in equity from Period 1 (aOR 0.61, 95% CI 0.55-0.67; **Figure 2D**) to Period 2 (aOR 0.77, 95% CI 0.73-0.81; **Figure 2D**), with Period 2 vs. Period 1 aOR of 2.97 (95% CI 2.65-3.33; **Table 2**).

Compared to those treated at rural hospitals, those treated at urban non-teaching and urban teaching hospitals were more likely to receive IVT in Period 1 (urban non-teaching vs. rural: aOR 1.80, 95% CI 1.52-2.12; urban teaching vs rural: aOR 3.17, 95% CI 2.65-3.80)**(Figure 2E**). Over time, IVT use increased in both rural and urban hospitals, but at a higher rate in urban hospitals, thus increasing the disparities in Period 2 (urban non-teaching vs. rural: aOR 2.58, 95% CI 2.33-2.85; urban teaching vs. rural: aOR 3.90, 95% CI 3.55-4.28; **Figure 2E**), with Period 2 vs. Period 1 aORs of 2.90 (95% CI 2.66-3.16) and 2.48 (95% CI 2.23-2.76) for urban non-teaching and urban teaching hospitals, respectively (**Table 2**).

**Figure 2D.**
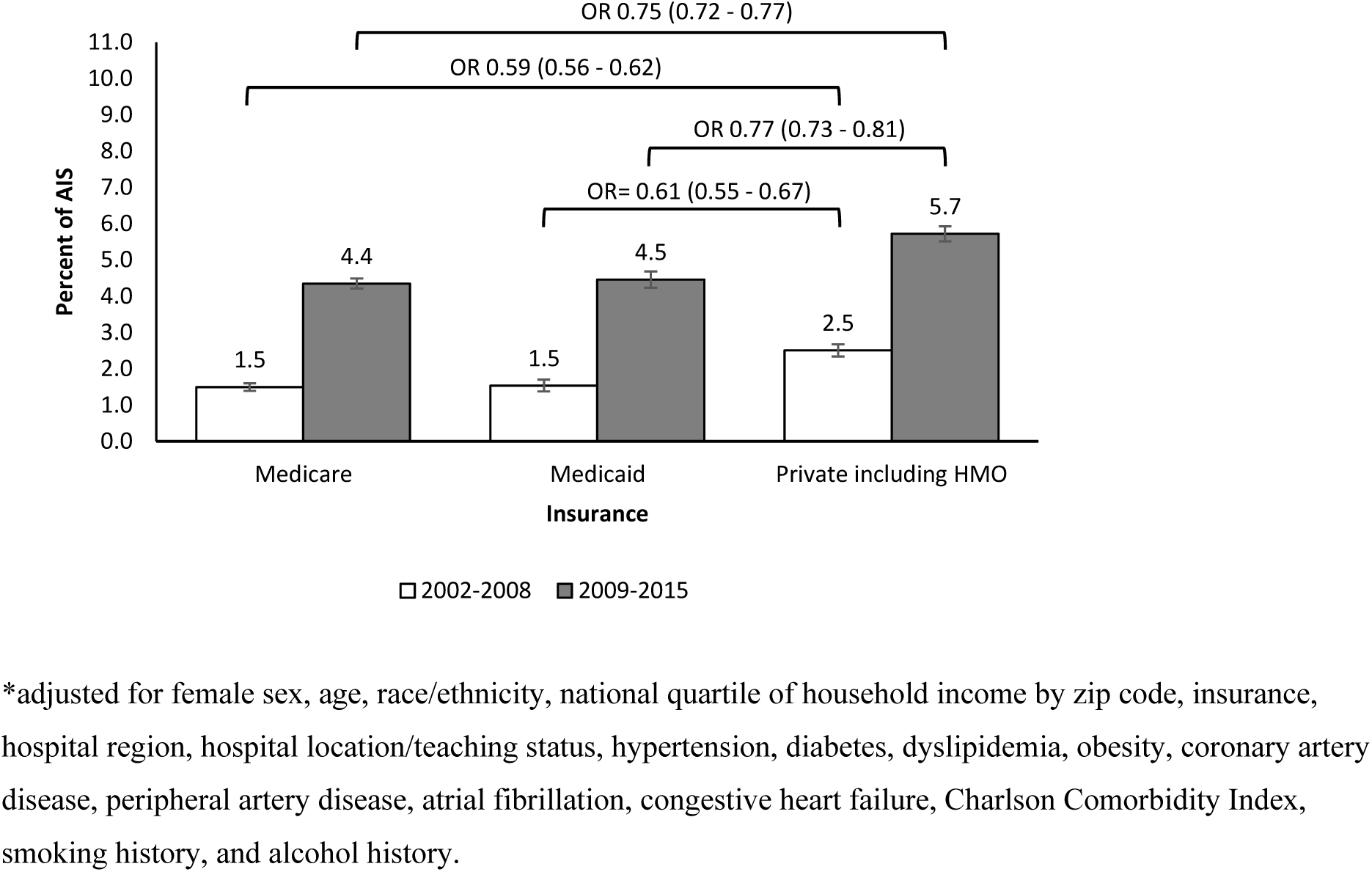
Dichotomized comparison of IVT use between *Period 1* vs. *Period 2* by insurance

**Figure 2E.**
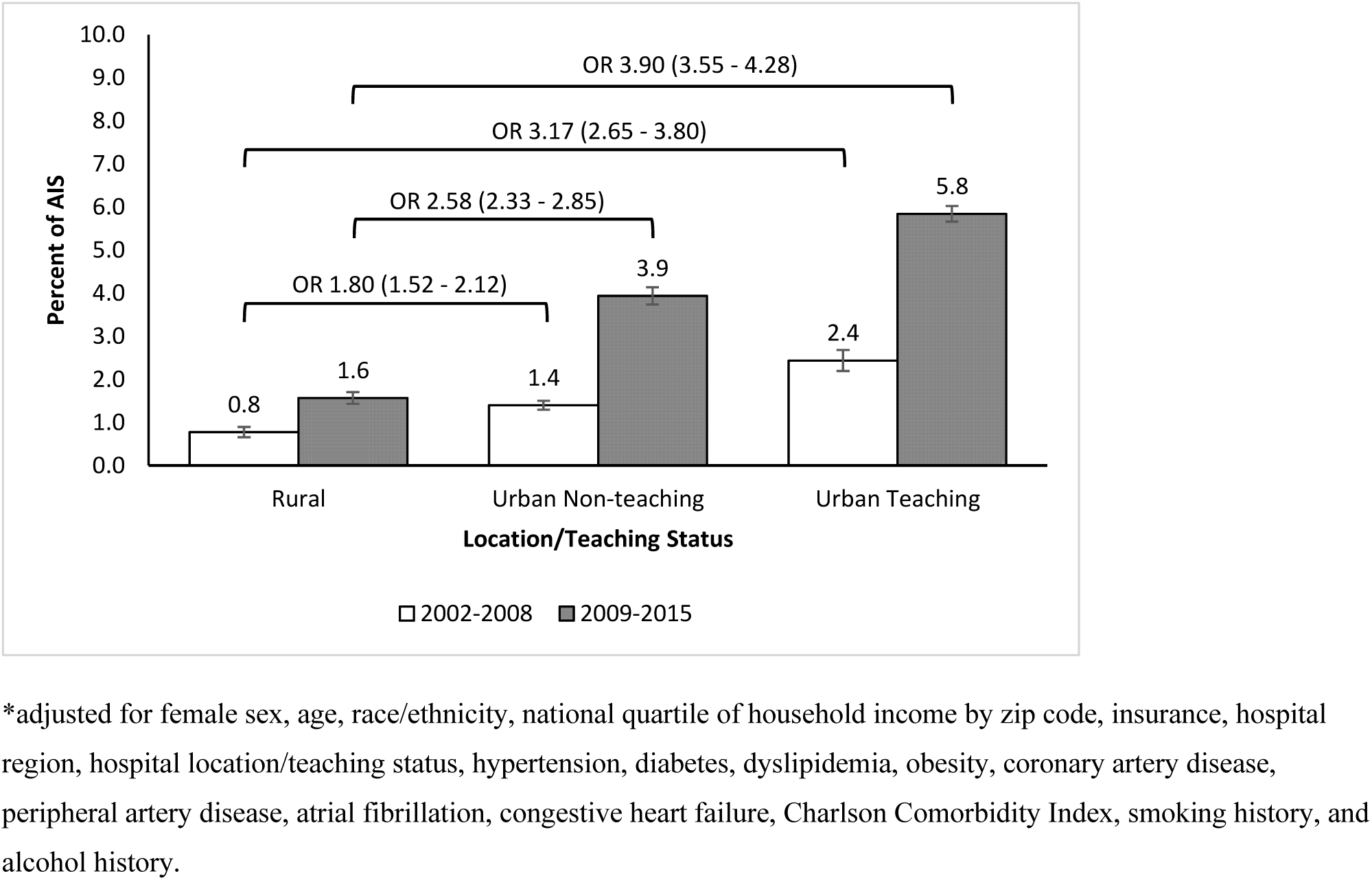
Dichotomized comparison of IVT use between *Period 1* vs. *Period 2* by hospital location/teaching status

## Discussion

This is the first study to assess temporal trends in IVT in the United States by numerous key sociodemographic and geographic characteristics, including race/ethnicity, sex, age, insurance, hospital teaching status, and geographic location. This study confirms steadily increasing IVT use across all ages in adults, with the fastest growth in those over 85 years. The sex inequity in IVT improved to near complete resolution in the second half of the study period. Inequities in IVT for Hispanic individuals resolved. Marked inequities in IVT remain for Black individuals despite improvements over time. Individuals with Medicare and Medicaid insurance remained less likely to receive IVT compared to privately-insured individuals, despite modest improvements over time. The inequities by geographic location continued to worsen, such that those admitted to rural hospitals were nearly three times less likely to receive IVT compared to those admitted to urban non-teaching hospitals and nearly 4 times less likely to receive IVT compared to those admitted to urban teaching hospitals, a finding consistent with recently published data^9^.

Individuals with AIS ≥85 years had the steepest relative growth in IVT. This corroborates a previous study examining trends from 2005 to 2010^3^, where individuals ≥85 years showed the most rapid increase in IVT, mostly in urban and high-volume hospitals. Similarly, a study using Get With the Guidelines-Stroke data^14^ and a recent study in Austria^15^ from their national Stroke Unit Registry noted a similarly dramatic increase in patients > 85 years of age and > 80 years.

Despite the increase in IVT, its absolute rates of use remained low in this age group. It is unclear whether this is due to patients presenting outside the therapeutic window, inability to determine last known well time in those who live alone, concerns about adverse events, or contraindications due to comorbidities. The under-utilization of IVT in individuals under the age of 18 is likely due to limited evidence of efficacy in this age group, lack of FDA-approval, atypical presentations, and lower index of suspicion for stroke.

This is the first study to show resolution of disparities in IVT for Hispanic individuals. However, race inequities remain. This study corroborates a previous NIS study that showed that Black individuals were less likely to receive IVT than White individuals^7,8^. A previous study using the NIS database in 2004-2010 noted under-administration in Black patients regardless of presentation to primary stroke centers.^8^ Potential reasons for the marked disparity for Black individuals could include provider implicit bias or discrimination,^16^ or longer time to presentation from factors such as poor access to care,^17,18^ medical mistrust^19^ and differences in stroke preparedness (i.e. the ability to recognize signs and symptoms of stroke, knowledge to call 911, and action to call 911).^20,21^ It could also be due to systematic differences in quality of care at hospitals where Black patients most often present.^7^

This study is the first to show an improvement in sex disparities in IVT. It is widely recognized that women are less likely to receive IVT compared to men. Factors contributing to sex differences in thrombolytic rates for women include delays in presentation^22^, atypical presentations^23^, underlying system-level factors^7^, inability to determine last known well time, and provider bias^24^. More research is needed to determine what has led to the reductions in sex differences, but more widespread use of stroke pathways may be a factor.

This study is the first to evaluate nationwide temporal trends in receipt of IVT by insurance type. The profound differences in IVT use by insurance type highlight the need to expand healthcare access and improve quality of care for those with government insurance. An example of an initiative that was successful in augmenting use of IVT was Target: Stroke quality initiative^25^, which led to a quicker administration of IVT with better long-term outcomes in Medicare beneficiaries.

This study confirms a persistent trend of worsening rural-urban disparities in IVT for AIS in the early 21^st^ century^4,6,9^. This widening disparity occurred with the steady temporal growth in IVT use in urban teaching hospitals; use of IVT in rural hospitals fluctuated in the young AIS patients aged 19-44 from 2001 to 2009^6^. A study from 2012 to 2017 demonstrated a persistent, steady gap in IVT use for rural populations^9^. The widening urban/rural gap could be explained by poor hospital and emergency medical services staffing, access to specialists, long distance to stroke centers, and stroke literacy. These issues could be mitigated by expansion of telestroke networks^26^ and community outreach.

These sociodemographic and geographic inequities are likely due to individual, system, and societal factors^3,4,6–9,27^; therefore a multipronged approach is needed to address them. Barriers to elimination of inequities include: (1) fundamental drivers of inequities, namely unequal distribution of wealth, education, and employment opportunities; (2) historical and ongoing structural and systemic racism which have disproportionately burdened Black communities with poverty, food insecurity, housing instability, and other adverse social determinants of health, and led to the likelihood of Black individuals having poor access to care^18,28^ or receiving care in under-resourced, under-performing hospitals; (3) ineffective messaging around stroke symptoms in lower income, Black, and Hispanic populations^20,21^; (4) provider-level factors, such as unconscious bias^29^, racism^30^, and hesitancy to treat the elderly with IVT;^3^ (5) patient-level factors, such as women being more likely to live alone^22^ and delays in presentation^17^, and (6) clinical factors such as atypical clinical presentations^23^.

The study is limited by its cross-sectional design and lack of patient-level zip codes and stroke specific data such as hospital stroke center designation, last known well times, stroke severity, and individualized considerations for IVT use (e.g., personal/goals of care decisions and medical contraindications not addressed by the exclusion criteria). Additionally, administrative data (e.g., race/ethnicity, diagnosis) are prone to misclassification and coding errors. We excluded individuals who were transferred in, so we may have underestimated IVT rates.

However, hospitals usually provide IVT prior to transfer, so we suspect these numbers are low. The strengths of this study include that it is nationally representative, with key hospital-level factors, sociodemographic characteristics, and comorbidities.

Further studies are needed to develop an understanding of reasons underlying persistent inequities as well as recent improvements (e.g., resolution in inequities faced by Hispanic individuals). It will be critical to elucidate the extent to which these inequities are caused by system-, provider-, society-, and patient-level factors. Developing a more nuanced understanding of the causes for persistent inequities in IVT will inform the development of effective interventions for reducing them.

## Conclusions

From 2002 through 2015, IVT for AIS in the United States increased steadily in various strata, with some encouraging trends of rapidly growing use of IVT among individuals ≥85 years and a resolution of disparities for Hispanic individuals. Despite these encouraging trends, only 1 in 15 AIS patients received IVT, and inequities remain for Black patients, women, those with Medicaid or Medicare insurance, and individuals admitted to rural hospitals. Further studies can help us better understand these trends and design interventions aimed at eliminating inequities in IVT for AIS.

## Data Availability

The raw data supporting the conclusions of this article will be made available by the authors, without undue reservation.

